# Doxycycline post-exposure prophylaxis among men who have sex with men and transgender women in Belgium: awareness, use, and antimicrobial resistance concerns in a cross-sectional online survey

**DOI:** 10.1101/2024.07.25.24310975

**Authors:** Thibaut Vanbaelen, Anke Rotsaert, Irith De Baetselier, Tom Platteau, Bernadette Hensen, Thijs Reyniers, Chris Kenyon

## Abstract

**Objectives:** We aimed to assess the awareness, willingness to use, and actual use of doxycycline post-exposure prophylaxis (doxyPEP) among men who have sex with men (MSM) and transgender women (TGW) in Belgium. Additionally, we aimed to identify factors associated with doxyPEP use and concerns regarding antimicrobial resistance (AMR).

**Methods:** Cross-sectional online survey among MSM and TGW in Belgium in April 2024. Participants were recruited through sexual networking applications and social media of community-based organizations. Numerical variables were compared with Wilcoxon rank-sum test and categorical variables with chi-square or Fisher’s exact tests. Factors associated with doxyPEP use were assessed using logistic regression. Willingness to use doxyPEP and concerns about side-effects/AMR were assessed before and after presenting a brief paragraph on the potential effects of doxyPEP on AMR.

**Results:** A total of 875 individuals initiated the survey. Almost all identified as men (860/875, 98.3%) with a median age of 40 years (IQR 32-48). Among all respondents, 40.4% (n=352/875) had heard of doxyPEP and 9.4% (n=82/875) had used it, among whom the majority used it within the last six months (70/81, 86.4%). In multivariable logistic regression, doxyPEP use was associated with having had ≥1 STI in the past 12 months and engagement in chemsex.

About 80% of the participants initially reported being willing to use doxyPEP, and about 50% reported being concerned about side effects. After reading about the potential effects of doxyPEP on AMR, willingness to use doxyPEP decreased to 60% and concerns of side-effects including AMR increased to around 70%.

**Conclusions:** Approximately one in ten MSM and TGW in Belgium reported using doxyPEP, with those at highest STI risk reporting higher usage. Importantly, concerns about AMR and side effect influenced willingness to use doxyPEP. If doxyPEP is introduced, informing patients about doxyPEP benefits and risks is crucial to enable informed decision-making.

**What is already known on this topic:** Several RCTs have shown the efficacy of doxycycline post-exposure prophylaxis (doxyPEP) on the incidence of chlamydia, syphilis, and in some instances gonorrhoea, among men who have sex with men (MSM) and transgender women (TGW). However, the potential for antimicrobial resistance (AMR) due to increased doxycycline consumption is a major concern, leading to some guidelines not recommending doxyPEP. Informal use of doxyPEP has been reported by up to 10% of MSM in countries where it is not recommended.

**What this study adds:** We found that about one in ten MSM in Belgium has ever used doxyPEP, with a majority having used it in the past six months. DoxyPEP use was associated with higher odds of having had one or more STIs in the preceding year and having engaged in chemsex in the past six months. The willingness to use doxyPEP was high but decreased after presenting information about the potential effects of doxyPEP on AMR. In contrast, concerns regarding doxyPEP side-effects were high and further increased after presenting information about the potential effects of doxyPEP on AMR.

**How this study might affect research, practice or policy:** By highlighting the prevalence and factors associated with informal doxyPEP use, this study can inform future research directions, guiding further investigations into patterns of STI prevention among MSM and TGW in Belgium. The insights the study adds of the impact of AMR concerns on the willingness to use doxyPEP, can influence clinical practice by emphasizing the importance of comprehensive patient education to ensure informed decision-making regarding STI prevention strategies. From a policy perspective, the study underscores the need for a comprehensive assessment of the challenges and benefits of doxyPEP, balancing its potential for reducing STI incidence with the risks of promoting antimicrobial resistance.

## Introduction

Three randomized controlled trials (RCT) have demonstrated the efficacy of doxycycline post-exposure prophylaxis (doxyPEP) in reducing the incidence of bacterial sexually transmitted infections (STIs) in men who have sex with men (MSM) (1–3). In these RCTs, doxyPEP reduced the incidence of *Chlamydia trachomatis* (CT) by 70-88% and syphilis by 73-87% (1, 2). The efficacy of doxyPEP on the incidence of *Neisseria gonorrhoeae* (NG) is less clear, with some studies demonstrating a reduction in NG cases, whereas others found no effect on incidence (2, 4). The main concerns related to doxyPEP use are that it could induce resistance to tetracyclines and other antimicrobials in NG and other bacteria, and could lead to deleterious alterations in the resistome and microbiome (5). For these reasons, while some guidelines promote the use of doxyPEP in MSM, others advised against it (6, 7).

There are reports of informal use of antibiotics for STI prevention in countries where doxyPEP is not currently recommended. For instance, in 2020, it has been shown that 2-10% of MSM reported using doxyPEP in the Netherlands, the United Kingdom (UK), and Australia (8–10). In Belgium, we found that about 3.2% of HIV pre-exposure prophylaxis (PrEP) users reported having ever taken antibiotics preventatively for bacterial STIs in 2022 (11). While the use seemed rather limited, the awareness of such practices was high, with about one third of PrEP users having heard of doxyPEP and one tenth knowing someone who currently used doxyPEP (11). DoxyPEP has also gained a lot of attention in the past year, following the publication of the results of several doxyPEP RCTs and its roll-out in San Francisco (12). We therefore hypothesized that the awareness and use of doxyPEP in Belgium has increased since 2022.

We aimed to assess the awareness, willingness to use, and use of doxyPEP among MSM and TGW in Belgium in 2024. Moreover, we aimed to assess socio-demographic factors and sexual behaviours associated with doxyPEP use and concerns regarding the effect of doxyPEP on AMR.

## Methods

### Study design and participants

We conducted a cross-sectional online survey among MSM and TGW in Belgium in April 2024. Participants were recruited through sexual networking applications (Grindr, Recon, Scruff, Jack’d) and social media platforms of community-based organizations. Potential participants were informed about the study and provided consent by agreeing to participate. Inclusion criteria were being 18 years old or older; assigned male sex at birth; identify as MSM or as a TGW; living in Belgium; had sex with at least one non-steady partner in the previous 12 months; being able to read and understand Dutch, French, or English; and willing and be able to provide informed consent. After eligibility check, eligible participants were invited to start the questionnaire.

The questionnaire was pilot tested by research team members and available in Dutch, French, or English. It included questions on socio-demographic characteristics (e.g., age, region of residence, highest level of education attained), awareness, willingness to use, current and past use of doxyPEP, sexual behaviours (e.g., engagement in chemsex, number of non-steady sexual partners, condom and PrEP use), and history of STI diagnoses (Appendix 1). Responses were collected anonymously, and participants were allowed to skip questions. Hence, the denominator of the collected answers may vary.

In the questionnaire, we first presented a short text describing doxyPEP and assessed awareness of doxyPEP using the questions “before today, had you ever heard of doxyPEP as a way to prevent bacterial sexually transmitted infections?” with the response options “yes” or “no”. Subsequent doxyPEP use questions were only asked to participants who had ever heard of doxyPEP, these included: “have you ever used or are you currently using doxyPEP?”. Response options were: “Yes, I am using it now”, “Yes, I have used it but not anymore”, and “No”. Subsequent questions about how doxyPEP was used were only asked to participants who reported current or past doxyPEP use.

Willingness to use doxyPEP and concerns of its side-effects were explored among all respondents, whether or not they had heard of doxyPEP. Willingness to use doxyPEP was assessed with the question “would you be willing to use doxyPEP to limit the risk of getting an STI?” with “certainly not”, “probably not”, “undecided”, “probably yes”, and “certainly” as response options. Concerns regarding doxyPEP side-effects were assessed by the question “with your current knowledge, how unconcerned or concerned are you about potential short- and long-term side effects of doxyPEP?” with “not concerned at all”, “rather not concerned”, “nor concerned nor unconcerned”, “rather concerned”, and “very concerned” as possible response options. Subsequently, we presented a short paragraph describing the potential effects of doxyPEP on AMR and assessed concerns of AMR with the question “with this additional information in mind, how unconcerned or concerned are you about doxyPEP leading to the emergence of antimicrobial resistance in sexually transmitted infections (STI)?” with the same possible response options as for the previous question. Lastly, we re-assessed the willingness to use doxyPEP using the same question as before.

### Data analysis

We described numerical variables using medians and interquartile ranges (IQR), and categorical variables using absolute numbers and proportions. Numerical variables were compared with Wilcoxon rank-sum test and categorical variables with chi-square test or Fisher’s exact test.

Willingness to use doxyPEP and concerns of side-effects/AMR before and after the short paragraph about AMR were compared with McNemar-Bowker test.

We explored whether socio-demographic characteristics and sexual behaviours were associated with doxyPEP use using logistic regression analyses. For this purpose, the variable doxyPEP use was dichotomized into participants who never took doxyPEP and participants reporting current or past doxyPEP use. We first performed univariable logistic regression to select the variables to include in a multivariable logistic regression analysis. Variables associated with doxyPEP use at the p≤0.10-level were included in a multivariable regression model. The final multivariable model was built by backward selection, using the likelihood ratio test and a significance level set at 0.05. The multivariable regression model was adjusted for age.

All computations were performed using R studio version 4.2.0 (13).

The STROBE checklist for reporting cross-sectional studies can be found in Appendix 2.

### Ethics approval

We obtained ethical approval from the Institutional Review Board of the Institute of Tropical Medicine (IRB 1753/24). All participants provided consent before participation in the study. Participation was anonymous.

## Results

### Survey participation and socio-demographic characteristics

A total of 1131 individuals accessed the survey between April 9 and April 30, 2024 and 948 (83.8%) agreed to participate. Among them, 73 did not meet the inclusion criteria and 875 started the survey. The median age of participants was 40 years (IQR 32-48, Table 1). Almost all participants identified as men (860/875, 98.3%), one identified as trans-women (1/875, 0.1%), twelve as non-binary (12/875, 1.4%), one as queer (1/875, 0.1%), and one as bisexual-gay (1/875, 0.1%). Most participants were born in Belgium (632/875, 72.2%), had completed or were completing short- or long-type higher education (725/875, 82.9%), had social security (838/875, 95.8%), and half lived in Flanders (501/875, 57.3%).

**Table 1.**
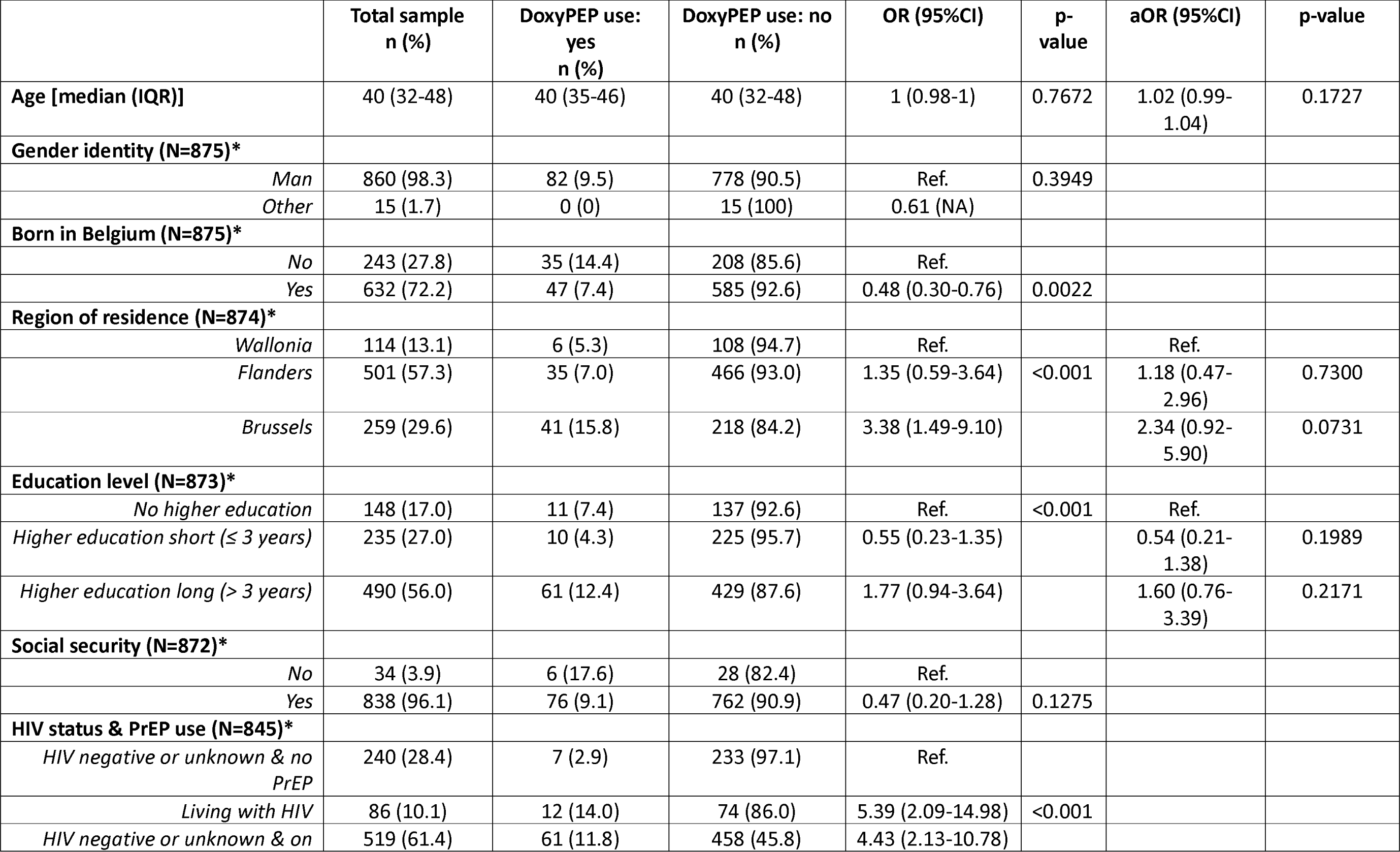
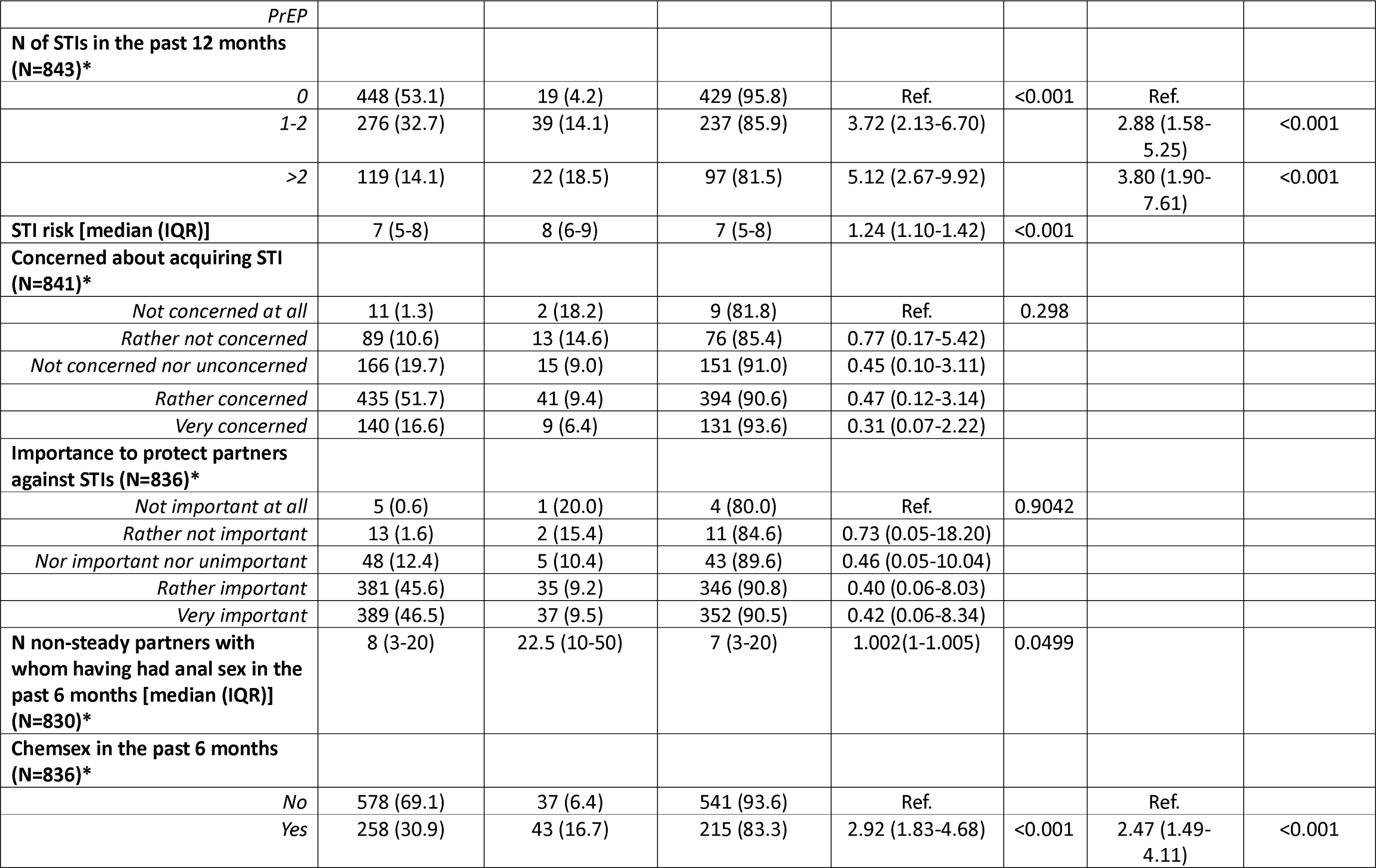
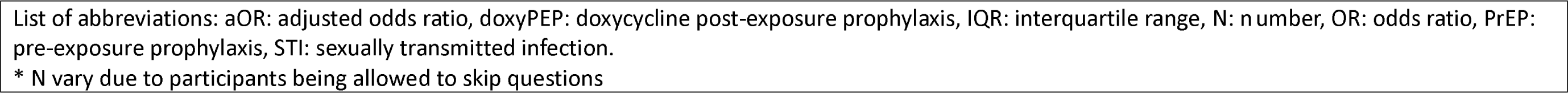
Factors associated with doxyPEP use in a univariable and multivariable logistic regression analysis.

### Sexual behaviours, HIV status and history of STIs

Eighty-six participants (86/845, 10.2%) were living with HIV, two-thirds (519/845, 61.4%), were HIV negative or had an unknown HIV status and were taking PrEP, and 240 were HIV negative or had an unknown HIV status and were not taking HIV-PrEP (240/845, 28.4%; Table 1). About half the participants reported an STI in the past year (395/843, 46.9%), and 119 had more than two STIs in the past year (119/843, 14.1%). The most frequently reported STIs were gonorrhoea (242/844, 28.7%), chlamydia (222/844, 26.3%), followed by syphilis (118/844, 14.0%). The median self-perceived STI risk was 7 (IQR 5-8) on a scale of 0 to 10 (0 representing very low risk and 10 very high risk). Most participants were rather or very concerned about acquiring STIs (575/841, 68.4%) and found it rather or very important to protect themselves or their partners against STIs (751/837, 89.7% and 770/836, 92.1%, respectively). Around one third of participants reported having engaged in chemsex in the previous 6 months (258/836, 30.9%).

### DoxyPEP awareness and use

About 40% of the participants had ever heard of doxyPEP as a way to prevent bacterial STIs (352/875, 40.2%, Table 2). Of these, around half had heard of it through community organizations (186/350, 53.1%). Interestingly, nine participants (9/350, 2.6%) mentioned in free text that they had heard about doxyPEP via international friends or organizations. About 10% of all participants had ever used doxyPEP (82/875, 9.4%). Among those, almost all took 200mg of doxycycline after sex (70/81, 86.4%), the majority had used it last in the previous six months (70/81, 86.4%), and used it less than monthly (51/81, 63%). DoxyPEP was mostly used when having group sex (60/80, 75%), when having sex with an anonymous partner (59/80, 73.8%) or when engaging in chemsex (33/80, 41.3%). DoxyPEP was used more frequently for receptive or insertive anal sex without a condom (62/80, 77.6% and 58/80, 72.5%, respectively) than for receptive or insertive oral sex without a condom (39/80, 48.8% and 43/80, 53.8%, respectively). The main sources of doxyPEP were leftover antibiotics (30/80, 37.5%) and getting doxycycline from friends or sex partners (23/80, 28.8%).

**Table 2.** DoxyPEP awareness and use.

|  | N respondents (%) |
| --- | --- |
| **Before today, had you ever heard of doxyPEP as a way to prevent bacterial sexually transmitted infections? (N=871)** |  |
| *Yes* | 352 (40.4) |
| *No* | 519 (59.6) |
| **How did you hear about doxyPEP?\* (N=350)** |  |
| *Community organizations* | 186 (53.1) |
| *Friends* | 43 (12.3) |
| *Healthcare professional* | 69 (19.7) |
| *Other* | 31 (8.9) |
| **Have you ever used or are you currently using doxyPEP? (N=350)** |  |
| *Yes, I am using it now* | 36 (10.3) |
| *Yes, I have used it but not anymore* | 46 (13.1) |
| *No* | 268 (76.6) |
| **How did you use or are you currently using doxyPEP?\* (N=81)** |  |
| *200mg of doxycycline after sex* | 70 (86.4) |
| *100mg daily* | 4 (4.9) |
| *200mg daily* | 5 (6.2) |
| *Other* | 4 (4.9) |
| **When did you last use doxyPEP? (N=81)** |  |
| *In the past month* | 39 (48.1) |
| *One to six months ago* | 31 (38.3) |
| *Seven to twelve months ago* | 6 (7.4) |
| *More than twelve months ago* | 5 (6.2) |
| **In the past 12 months, how often did you take doxyPEP? (N=81)** |  |
| *Daily or almost daily* | 3 (3.7) |
| *Weekly* | 6 (7.4) |
| *Monthly* | 21 (25.9) |
| *Less than monthly* | 51 (63.0) |
| **In which of the following situations did you use doxyPEP?\* (N=80)** |  |
| *When having sex with a steady partner* | 3 (37.5) |
| *When having sex with a regular casual sex partner* | 14 (17.5) |
| *When having sex with an anonymous partner* | 59 (73.8) |
| *When having group sex* | 60 (75.0) |
| *When combining drugs and sex ("chemsex")* | 33 (41.3) |
| *Other* | 4 (5.0) |
| **In which of the following situations did you use doxyPEP?\* (N=80)** |  |
| *Oral receptive sex without a condom* | 43 (53.8) |
| *Oral insertive sex without a condom* | 39 (48.8) |
| *Anal receptive sex without a condom* | 62 (77.6) |
| *Anal insertive sex without a condom* | 58 (72.5) |
| *Oral receptive sex with a condom* | 3 (3.8) |
| *Oral insertive sex with a condom* | 2 (2.5) |
| *Anal receptive sex with a condom* | 6 (7.5) |
| *Anal insertive sex with a condom* | 4 (5.0) |
| **How did you obtain doxyPEP?\* (N=80)** |  |
| *My GP prescribed it for me* | 10 (12.5) |
| *A doctor in an HIV/STI/PrEP clinic prescribed it to me* | 14 (17.5) |
| *Another healthcare professional prescribed it to me* | 11 (13.8) |
| *I used antibiotics that are left over from a different treatment* | 30 (37.5) |
| *I bought it online* | 11 (13.8) |
| *I got it from friends/sex partners* | 23 (28.8) |
| *Other (specify)* | 9 (11.3) |
| * multiple answers possible<br>List of abbreviations: doxyPEP: doxycycline post-exposure prophylaxis, GP: general practitioner, HIV: human immunodeficiency virus, PrEP: pre-exposure prophylaxis, STI: sexually transmitted infection. |  |

### Factors associated with doxyPEP use

In univariable logistic regression analyses, current or past doxyPEP users were more likely to reside in Brussels or in Flanders than in Wallonia (OR 3.38 [95%CI 1.49-9.10] and OR 1.35 [95%CI 0.59-3.64], respectively). They were also more likely to have completed or be in higher education compared with having no higher education (OR 1.77 [95%CI 0.94-3.64]), and to be living with HIV or being HIV negative or unknown HIV status and taking PrEP compared with being HIV negative or unknown HIV status and not taking PrEP (OR 5.39 [95%CI 2.09-14.98] and OR 4.43 [95%CI 2.13-10.78], respectively). DoxyPEP use was associated with having had 1-2 or >2 STIs in the past year (OR 3.72 [95%CI 2.13-6.70], and OR 5.12 [95%CI 2.67-9.92[, respectively), self-perceived a higher STI risk (OR 1.24 [95%CI 1.10-1.42]), having had a higher number of non-steady partners for anal sex in the past 6 months (OR 1.002 [95%CI 1-1.005]), and having engaged in chemsex in the past 6 months (OR 2.92 [95%CI 1.83-4.68]).

In the multivariable regression analysis, doxyPEP use remained significantly associated with having had 1-2 or >2 STIs in the past 12 months (OR 2.88 [95%CI 1.58-5.25] and OR 3.80 [95%CI 1.90-7.61], respectively), and engagement in chemsex in the past 6 months (OR 2.47 [95%CI 1.49-4.11]), while adjusting for age.

### Willingness to use doxyPEP and concerns of side-effects and AMR

About 80% of the participants initially reported being certainly or probably willing to use doxyPEP (387/868, 44.6% and 331/868, 38.1%, respectively, Figure 1), and about 7% were probably or certainly not willing to use it (50/868, 5.8% and 7/868, 0.8%, respectively). After reading a paragraph describing the potential effects of doxyPEP on AMR, the proportion of those being certainly or probably willing to use doxyPEP declined to about 60% (193/859, 22.5% and 327/859, 38.1%, respectively) and the proportion of those probably or certainly not willing to use it increased to about 18% (125/859, 14.6% and 31/859, 3.6%, respectively). This change was statistically significant (p<0.001).

**Figure 1.**
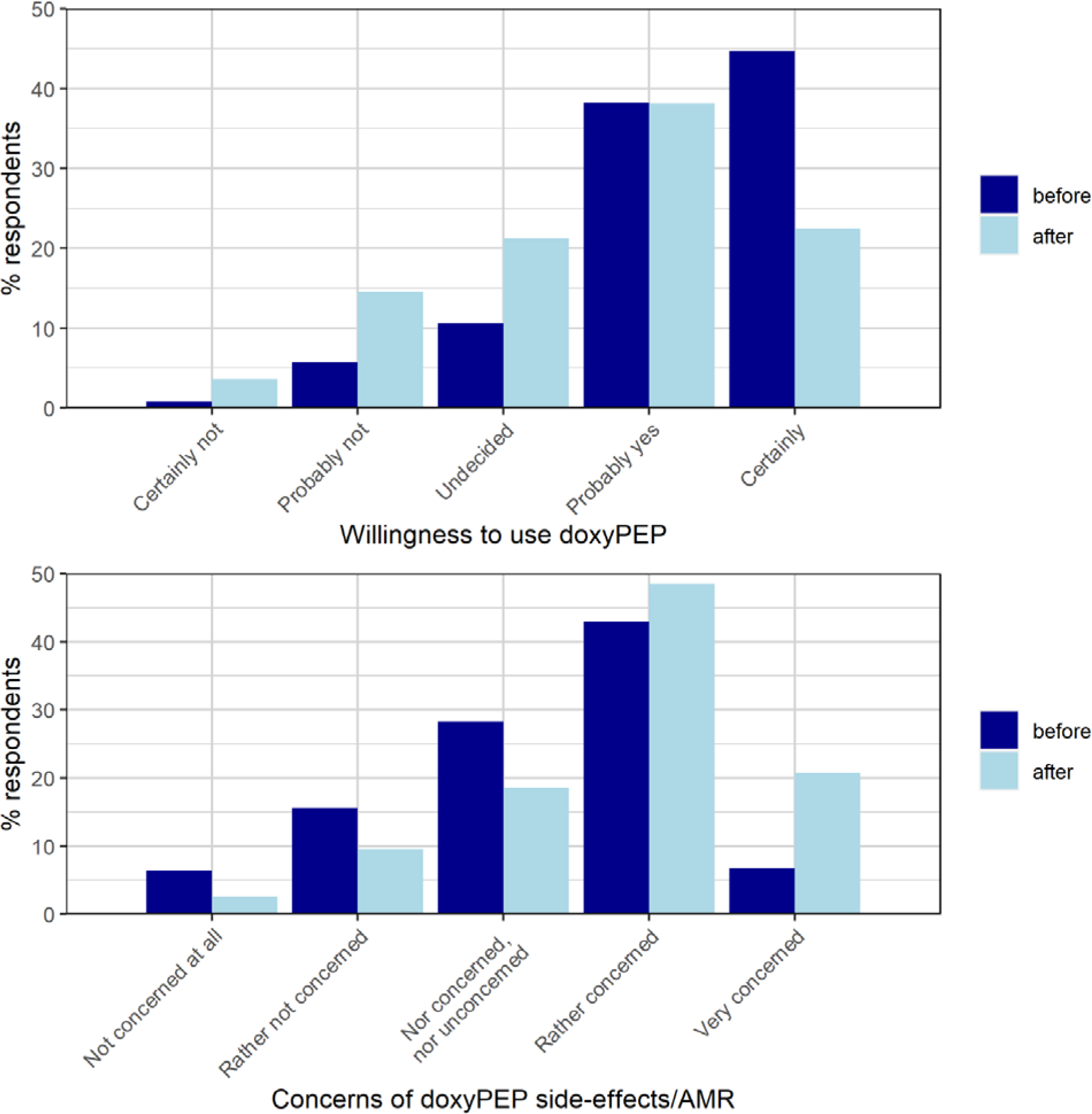
Willingness to use doxyPEP (above panel) and concerns of side-effects and AMR (below panel). AMR: antimicrobial resistance, doxyPEP: doxycycline post-exposure prophylaxis.

About half of the participants initially reported being very or rather concerned about potential short- and long-term side effects of doxyPEP (58/859, 6.7% and 371/859, 42.7%, respectively, Figure 1), and about a quarter were rather not or not at all concerned (135/859, 15.6%, and 55/859, 6.4%, respectively). After reading the short AMR paragraph, more than two thirds of the participants were rather or very concerned about the potential for doxyPEP leading to AMR (178/859, 20.7% and 417/859, 48.5%, respectively), and about one in ten were rather not or not at all concerned (82/859, 9.5%, and 22/859, 2.6%, respectively).

## Discussion

We found that about one in ten MSM in Belgium has ever used doxyPEP, with a majority of these individuals having used it the prior six months. DoxyPEP use was associated with having had STIs in the preceding year having engaged in chemsex in the preceding 6 months. Willingness to use doxyPEP was high but decreased after presenting information about the potential effects of doxyPEP on AMR while concerns of doxyPEP side-effects increased after presenting information about the potential effects of doxyPEP on AMR.

Our finding that approximately 10% of MSM in Belgium have used doxyPEP is consistent with previous studies. Surveys in the UK and Australia found that 9% and 10% of PrEP users were already using doxyPEP, respectively, despite not being formally recommended at the time (10, 14). In Belgium in 2022, we found that 3.2% of PrEP users were using antibiotics for STI prevention and 28.9% had heard of it (11). These proportions seem to have increased, although careful consideration should be taken when comparing studies with different sampling methodologies, timeframes, and study populations. The publication of doxyPEP RCT results and its implementation in San Francisco may have increased awareness and informal use among MSM. Some participants in our study reported learning about doxyPEP through international friends or organizations, supporting this hypothesis. Furthermore, other European countries recently reported higher levels of informal doxyPEP use. In Germany, 29% of MSM reported ever having used doxyPEP in a survey in 2023 (15).

Consistent with previous studies, we found that doxyPEP use was associated with engagement in chemsex, PrEP use, and recent STIs (10, 16, 17). It has been shown that chemsex and PrEP use are associated with higher rates of STIs (18, 19). Our findings suggest that those who are the most at risk for STIs are the ones reporting doxyPEP use. Moreover, among doxyPEP users the main circumstances of use were high-risk events for STIs like group sex or sex under the influence of substances. While it might seem appropriate that those who are the most at risk for STIs are the ones using doxyPEP, careful consideration should be taken to high antimicrobial consumption in this population. Previous studies have shown that AMR has frequently emerged in core-groups with high antimicrobial consumption, before spreading to other populations (20). We, and others, have previously shown that rolling out doxyPEP would lead to a substantial increase in doxycycline consumption, leading to a risk of selecting or inducing antimicrobial resistance in a wide-range of pathogens, including *Neisseria gonorrhoeae* (21–25). While doxyPEP may have the largest STI incidence impact in high-risk groups, it may also have the greatest AMR effect in this group.

After providing information about the potential risks of AMR associated with doxyPEP, willingness to use doxyPEP decreased and the concerns of side-effects/AMR increased. Previous qualitative studies reported high willingness to use doxyPEP despite up to 50% of participants voicing AMR concerns (26, 27). In these studies, participants reported that more information about the potential side-effects of doxyPEP, including AMR, is necessary before deciding on doxyPEP use. Our study is the first to assess how providing AMR information can influence the willingness to use doxyPEP. These insights underscore the importance of informing potential users about both the benefits and harms of doxyPEP to enable informed decisions about use.

Our study has several limitations. First, the study was advertised on a limited number of sexual networking applications and social media platforms. Moreover, potential self-selection is inherent to the online study design. Hence, the sample may not be representative of the entire MSM population in Belgium. Our survey was advertised as a doxyPEP survey, which might have led to individuals who knew doxyPEP being overrepresented in our sample. Second, participants responses might be affected by a recall bias and, given the sensitive and nature of the questions, subjects might be prone to social desirability bias, which could have led to underreporting of sexual risk taking. Lastly, since doxyPEP is currently not recommended in Belgium, participants may have been less willing to report informal use.

In conclusion, informal doxyPEP use appears to be increasing among MSM and TGW in Belgium, with those at highest STI risk reporting more use. While targeting doxyPEP to those at highest risk for STIs may seem appropriate, high antimicrobial consumption in this population raises AMR concerns. Importantly, AMR and side effect concerns could influence willingness to use doxyPEP. If doxyPEP is introduced, informing patients about the benefits and risks of doxyPEP will be crucial to enable informed decision-making.

## Data Availability

All data produced in the present study are available upon reasonable request to the authors.

