## Appendix 1 for "Doxycycline post-exposure prophylaxis among men who have sex with men and transgender women in Belgium: awareness, use, and antimicrobial resistance concerns in a cross-sectional online survey"

**Appendix 1 –Survey questionnaire**

**Eligibility questions**

1. How old are you?
   - Dropdown list
2. Do you live in Belgium?
   - Yes
   - No
3. Which sex were you assigned at birth?
   - Male
   - Female
4. What is your current gender identity?
   - Man
   - Woman
   - Trans woman (person with a male sex at birth, but a woman gender identity)
   - Trans man (person with a female sex at birth, but a man gender identity)
   - Non-binary gender
   - Other (Please specify)
5. Who are you sexually attracted to (multiple answers possible)?
   - Men
   - Women
   - Trans women (persons with a male sex at birth, but a woman gender identity)
   - Trans men (persons with a female sex at birth, but a man gender identity)
   - None of these
   - Other (Please specify)
6. Did you have sex with at least one non-steady partner in the previous 12 months?

With non-steady partner we mean someone else than your husband or wife or stable (girl/boy) friend. A non-steady sex partner can be a sex buddy, fuck friend, friend with benefits, anonymous sex partner, etc.

- Yes
- No

**Socio-demographics**

7. Were you born in Belgium?
   - Yes
   - No
8. In which country were you born?
   - Dropdown list
9. In which Belgian region do you currently live?
   - Flanders
   - Brussels
   - Wallonia
10. What is the highest level of education that you have completed or that you are currently completing?
    - None
    - Primary school
    - Secondary school
    - Higher education, short type (3 years or less)
    - Higher education, long type (more than 3 years)
    - Other (Please specify)
11. Do you have social health insurance? (such as Christelijke Mutualiteit - CM, SocMut, ...)
    - Yes
    - No

**DoxyPEP questions**

**Doxycycline post-exposure prophylaxis (doxyPEP) is the use of an antibiotic (doxycycline) after sex in order to avoid getting a bacterial sexually transmitted infection (STIs) such as chlamydia, or syphilis. Studies have shown that it reduces the occurrence of these infections by around 80% in men who have sex with men (MSM). The effect on gonorrhea is unclear. There is some evidence that daily intake of doxycycline preventively works as well. However, there are still open questions concerning the long-term consequences of the use of doxyPEP. As such, the use of doxyPEP is not implemented in regular care so far.**

12. Before today, had you ever heard of doxyPEP as a way to prevent bacterial sexually transmitted infections?
    - Yes
    - No
13. If yes, how did you hear about doxyPEP? (select al that apply)
    - Online
    - Community organizations
    - Friends
    - Healthcare professional
    - Other (Please specify)
14. Would you be willing to use doxyPEP to limit the risk of getting an STI?
    - Certainly not
    - Probably not
    - Undecided
    - Probably yes
    - Certainly
15. Have you ever used or are you currently using doxyPEP?
    - Yes, I am using it now
    - Yes, I have used it but not anymore
    - No
16. If yes, how did you use or are you currently using doxyPEP?
    - 200mg of doxycycline after sex (1 tablet of 200mg or 2 tablet of 100mg)
    - 100mg daily (1 tablet of 100mg of ½ tablet of 200mg)
    - 200mg daily (1 tablet of 200mg or 2 tablet of 100mg)
    - Other (specify):
17. If yes, when did you last use doxyPEP?
    - In the past month
    - One to six months ago
    - Seven to twelve months ago
    - More than twelve months ago
18. If yes, in the past 12 months, how often did you take doxyPEP?
    - Daily (at least once a day) or almost daily
    - Weekly (at least once a week)
    - Monthly (at least once a month)
    - Less than monthly (less than once a month)
19. If yes, in which of the following situations did you use doxyPEP? (Select all that apply)

- When having sex with a steady partner

*With steady partner we mean a husband or a wife or stable (girl) friend, that you are not single and consider yourself to have a serious relationship with. Length of the relationship does not matter.*

- When having sex with a regular casual sex partner

*With a regular casual sex partner, we mean a person with whom you have regular sex but not a steady relationship, but who is also not anonymous (e.g. fuck buddies, 'friends with benefits', sex buddies)*

- When having sex with an anonymous partner

*With an anonymous or new sex partner we mean a person who you don't really know or you just met. e.g. someone you meet for sex for the first time after online contact on Grindr*

- When having group sex
- When combining drugs and sex (“chemsex”)
- Other (specify)

20. If yes, in which of the following situations did you use doxyPEP? (Select all that apply)

- Oral receptive sex **without** a condom (“to give a blowjob”)
- Oral insertive sex **without** a condom (“to receive a blowjob”)
- Anal receptive sex **without** a condom (“bottom”)
- Anal insertive sex **without** a condom (“top”)
- Oral receptive sex **with** a condom (“to give a blowjob”)
- Oral insertive sex **with** a condom ("to receive a blowjob”)
- Anal receptive sex **with** a condom (“bottom”)
- Anal insertive sex **with** a condom (“top”)

21. If yes, how did you obtain doxyPEP?

- My GP prescribed it for me
- A doctor in an HIV/STI/PrEP clinic prescribed it to me
- Another healthcare professional prescribed it to me
- I used antibiotics that are left over from a different treatment
- I buy it online
- I get it from friends/sex partners
- Other (specify)

22. With your current knowledge, how unconcerned or concerned are you about potential short- and long-term side effects of doxyPEP?

- Not concerned at all
- Rather not concerned
- Nor concerned nor unconcerned
- Rather concerned
- Very concerned

**The main concern related to doxyPEP use is that it might make bacteria resistant to antibiotics (i.e. induce antimicrobial resistance). This means infections could become harder to treat with antibiotics. Other studies have also shown that doxyPEP may not prevent gonorrhea as this infection is already resistant to doxycycline.**

23. With this additional information in mind, how unconcerned or concerned are you about doxyPEP leading to the emergence of antimicrobial resistance in sexually transmitted infections (STI)?
    - Not concerned at all
    - Rather not concerned
    - Nor concerned nor unconcerned
    - Rather concerned
    - Very concerned
24. With this additional information in mind, would you be willing to use doxyPEP to limit the risk of getting an STI?
    - Certainly not
    - Probably not
    - Undecided
    - Probably yes
    - Certainly
25. Would you be willing to participate in a doxyPEP study in which there is a 50% chance of receiving a placebo?
    - Yes
    - No

**Sexual health and sexuality**

26. Did you ever do a test for HIV?
    - Yes
    - No
27. If yes, what was the result of your last HIV test?
    - Negative
    - Positive
    - I don't know
    - I prefer not to say
28. Have you ever heard or used HIV pre-exposure prophylaxis (PrEP)? PrEP is a way of preventing HIV infection, for more information click [here](https://www.exaequo.be/en/hauptnavigation/your-health/pep-prep).
    - Yes
    - No
29. If yes, in the past 12 months, have you ever used HIV PrEP?
    - Yes
    - No
30. In the past 12 months, did you have any sexually transmitted infection (e.g. chlamydia, gonorrhea, syphilis)?
    - Yes
    - No
31. If yes, in the past 12 months, which of the following sexually transmitted infections did you have? (select all that apply)
    - Gonorrhea
    - Chlamydia
    - LGV (Lymphogranuloma venereum)
    - Syphilis
    - Mycoplasma genitalium
    - Other (Please specify)
    - I don’t know
32. If Gonorrhea, how many times did you have Gonorrhea in the past 12 months?
    - Dropdown list (1, 2, 3, 4, 5, >5)
33. If Chlamydia, how many times did you have Chlamydia in the past 12 months?
    - Dropdown list (1, 2, 3, 4, 5, >5)
34. If LGV, how many times did you have LGV in the past 12 months?
    - Dropdown list (1, 2, 3, 4, 5, >5)
35. If *Mycoplasma genitalium*, how many times did you have Mycoplasma genitalium in the past 12 months?
    - Dropdown list (1, 2, 3, 4, 5, >5)
36. If Syphilis, how many times did you have Syphilis in the past 12 months?
    - Dropdown list (1, 2, 3, 4, 5, >5)
37. If other, how many times did you have this STI in the past 12 months?
    - Dropdown list (1, 2, 3, 4, 5, >5)
38. On a scale from 1 to 10 (1= very low risk, 10= very high risk), to what extent do you consider yourself to be at risk for acquiring a sexually transmitted infection (e.g. syphilis, gonorrhea, or chlamydia)?
    - Scale 1-10
39. How unconcerned or concerned are you about acquiring a sexually transmitted infection (STI)?
    - Not concerned at all
    - Rather not concerned
    - Not concerned nor unconcerned
    - Rather concerned
    - Very concerned
40. How unimportant or important is it for you to protect **yourself** against STIs?
    - Not important at all
    - Rather not important
    - Nor important nor unimportant
    - Rather important
    - Very important
41. How unimportant or important is it for you to protect **your sex partners** against STIs?
    - Not important at all
    - Rather not important
    - Nor important nor unimportant
    - Rather important
    - Very important
42. In the past six months, did you have anal sex with a non-steady partner?

*With non-steady partner we mean someone else than your husband or wife or stable (girl/boy) friend. A non-steady sex partner can be a sex buddy, fuck friend, friend with benefits, anonymous sex partner, etc.*

- Yes
- No

43. If yes, with how many non-steady sex partners did you have anal sex? (Give an approximate number if you don’t know the exact number)
    - Free text only numbers allowed
44. In the past six months, how frequently did you use condoms for anal sex with non-steady sexual partners?
    - Never
    - Sometimes
    - Half of the time
    - Most of the time
    - Always
    - I prefer not to answer
45. In the past six months, have you had sex under the influence of substances ("chemsex")?

*For more information about sex under the influence of substances (“chemsex”) and the risks associated with it, click* [*here*](https://www.exaequo.be/en/hauptnavigation/gay-life/chemsex)*.*

- Yes
- No

46. If yes, what substances have you used (multiple answers possible)?
    - Cathinones (3MMC, 4MMC, 2MMC,4MEC, 4CMC, ...)
    - Cocaine
    - Amphetamine ('speed')
    - Methamphetamine ('crystal meth', 'TINA')
    - MDMA / Ecstasy / XTC
    - 2C-B
    - Cannabis
    - Alcohol
    - GHB
    - GBL
    - Poppers
    - Ketamine
    - Synthetic cannabioids
    - α-PHP, α-PVP
    - Methylphenidate ('Rilatin')
    - Erectile enhancing medication ('Viagra,' 'Kamagra,' etc.)
    - Benzodiazepines (diazepam, Valium, etc.)
    - Fentanyl
    - Other (please specify)
47. How often were you under the influence of substances during sex?
    - Never
    - Sometimes
    - Half the time
    - Most of the time
    - Always
    - I prefer not to answer
48. During substance-induced sex, were personal boundaries always respected (multiple answers possible)?
    - Yes, my personal boundaries were always respected, and I always respected the personal boundaries of my sexual partner(s).
    - No, my personal boundaries were not always respected by my sexual partner(s)
    - No, I did not always respect the personal boundaries of my sexual partner(s)
49. Did your drug use during sex have a negative impact on other areas of your life (multiple answers possible)?
    - No
    - Yes, on my relationship
    - Yes, on my family life
    - Yes, on my friendships
    - Yes, on my professional life (work)
    - Yes, on my physical health
    - Yes, on my mental health
    - Yes, on my sexual health
    - Yes, on another domain, namely ... (free text)
